# Self-reported Stress Perception in Breast Cancer Patients Treated with Integrative Metal Color Light Therapy: The LUCIA Study Protocol

**DOI:** 10.1101/2024.12.17.24319155

**Authors:** Shiao Li Oei, Jessica Groß, Michaela Ganz, Charlotte Streffer, Gerrit Grieb, Lysander Freytag, Sophia Johnson, Friedemann Schad, Anja Thronicke

## Abstract

**Introduction:** Many breast cancer patients benefit from appropriate oncological therapies, but also experience side effects or ongoing restrictions that have a significant impact on health-related quality of life. Non-Pharmaceutical Interventions (NPIs) such as sport, movement, art therapy, yoga or mindfulness-based stress reduction, can help to reduce distress and pain and support resilience strategies. Metal Colored Light (MCL) therapy, an integrative art therapy using metal colored glasses, has been used in the past to relieve symptoms, improve quality of life and support coping strategies. The LUCIA study aims for the first time to systematically investigate whether self-reported perception of stress can be influenced in breast cancer patients through a series of MCL.

**Methods and analysis:** The LUCIA-study will enroll 60 breast cancer patients treated with MCL therapy, which consists of nine individual MCL sessions per patient over a six-week period. The primary outcome is the analysis of self-reported stress perception of breast cancer patients using the National Comprehensive Cancer Network (NCCN) Distress Thermometer. Secondary endpoints include the Internal Coherence Scale (ICS) and quality of life using the European Organization for Research and Treatment of Cancer Questionnaire C30 (EORTC-QLQ-C30). Longitudinal changes in symptom burden will be assessed and multivariate analyses performed.

**Ethics and disseminations:** The present study is approved by the ethics committee of the Medical Association Berlin within the Network Oncology with the reference number Eth-27/10. The results will be presented at scientific conferences and published.

**Trial registration number:** The study was registered at the German Register for Clinical Trials under DRKS00013335 on 27/11/2017.

**Strengths and limitations of this study:**

- Self-reported stress perception of each participant in this LUCIA study will be evaluated before (T0) and after the Metal Colored Light (MCL) therapy (T1).
- Changes in self-reported distress burden, internal coherence and health-related quality of life will be followed and asked three months (T2) and six months (T3) after T0.
- Prospective evaluation of all participants.
- Sources of bias and potential confounding can be limited through the use of multivariate analyses.
- There is no direct control or placebo group and no blinding.

## Introduction

A growing number of people are living with cancer or have survived it [1]. In addition to advances in early detection and the increasing effectiveness of oncology therapies, the well-being of patients who have successfully survived cancer has become an increasingly important area of research. The majority of breast cancer patients suffer from persistent impairments after completion of their primary therapy. Oncology therapies are often associated with long-term effects. Long-lasting and late effects of cancer treatments can have a significant impact on patients’ distress and health-related quality of life, even years later [2]. The tumor itself, as well as the oncological therapies received, interferes with various metabolic pathways and hormonal regulatory circuits. This affects a variety of impairments such as chronic pain, cognitive impairment, and severe fatigue, known as cancer-related fatigue [3,4]. Experience with treatment programs for breast cancer patients has shown that, in addition to physical activity, conscious emotional management plays an important role in strengthening physical fitness and mental well-being [5,6]. Meditation and yoga in particular have been shown to be effective, music therapy can also help to reduce anxiety and stress [7], and dance therapy, such as Tango Argentino, can alleviate cancer-related fatigue in breast cancer survivors [8]. Art therapy is recognized as a mental health intervention with the goal of improving or restoring a person’s functioning and sense of personal well-being [9]. A study of adult women found that art therapy may be effective in treating depression and anxiety [10]. A systematic review provides some evidence that art therapy may help breast cancer patients with anxiety, depression, and fatigue [11]. Low level light therapy, also known as photobiomodulation therapy has been used for more than 40 years as a non-invasive treatment option to stimulate wound healing and reduce inflammation, edema and pain [12]. This light therapy uses non-ionizing light sources such as laser diodes in the visible and near-infrared spectrum and has emerged as a supportive care treatment modality for the management of breast cancer-related lymphedema, improving shoulder mobility and pain [13]. A meta-analysis of five randomized controlled trials involving a total of 231 participants with Parkinson’s disease evaluated the efficacy and safety of light therapy with active light sources [14]. In this meta-analysis, it was concluded that light therapy has a significant effect on both motor and non-motor function in patients with Parkinson’s disease. There was a significant improvement in depression and on sleep [14]. The basic mechanisms of light therapy are not yet fully understood; they could be based on the absorption of light by the body’s own chromophores, which can lead to physiological changes [15]. Light drives the melatonin metabolism, which influences the sleep-wake rhythm and regulates the circadian rhythm [16]. Unlike light therapy with artificial light sources, MCL therapy uses only natural daylight, shining through MCL glasses [17]. It was hypothesized, that MCL may stimulate the metabolism and promote mental balance to alleviate symptoms of stress, psychological strain and disease management [17].

The aim of the study is to analyze whether the perception of stress could be changed or even improved and if so, which of the stress-related impairments can be addressed by MCL.

## Methods

This report follows the Standard Protocol Items: Recommendations for Interventional Trials (SPIRIT) guidelines for the minimum content of a clinical trial protocol [18] (See Supplementary File 1 for the SPIRIT checklist).

### Study setting

This study is conducted by the Research Institute Havelhöhe (Forschungsinstitut Havelhöhe, FIH) at the hospital Gemeinschaftskrankenhaus Havelhöhe in Berlin (GKHB) within the Network Oncology (NO) [19]. The study participants are recruited at the primary care facility of the German Cancer Society – certified Breast Cancer Centre at the GKHB.

### Eligibility criteria

#### Inclusion and exclusion criteria for participants

Participants eligible for inclusion in the study must meet the following criteria:

- signed written informed consent and
- age ≥ 18 years and
- all stage breast cancer
- diagnosis of breast cancer 6 to 36 months before enrollment

Patients will be excluded when they meet one of the following exclusion criteria:

- no signed written informed consent
- pregnancy or breastfeeding
- concurrent participation in other clinical trials
- receiving concurrent chemotherapy or radiation therapy
- linguistic, medical, psychiatric, cognitive or other conditions that may compromise the patient’s ability to understand the patient information, comply with the study protocol or complete the study.

#### Who will take informed consent?

It is the responsibility of the study physician to provide sufficient verbal and written information on the study’s purpose and procedures, information on data protection procedures, possible advantages and disadvantages of participation, and on the option for the patient to withdraw from the study at any time and without any given reason and to take informed consent. Written informed consent will be obtained from all participants prior study enrolment.

### Interventions

#### Intervention description

The MCL therapy sessions take place in a darkened room; the only daylight in the room is that which passes through the MCL panels. The patient is seated at a distance of about one meter in front of the MCL panel (Figure 1).

**Figure 1.**
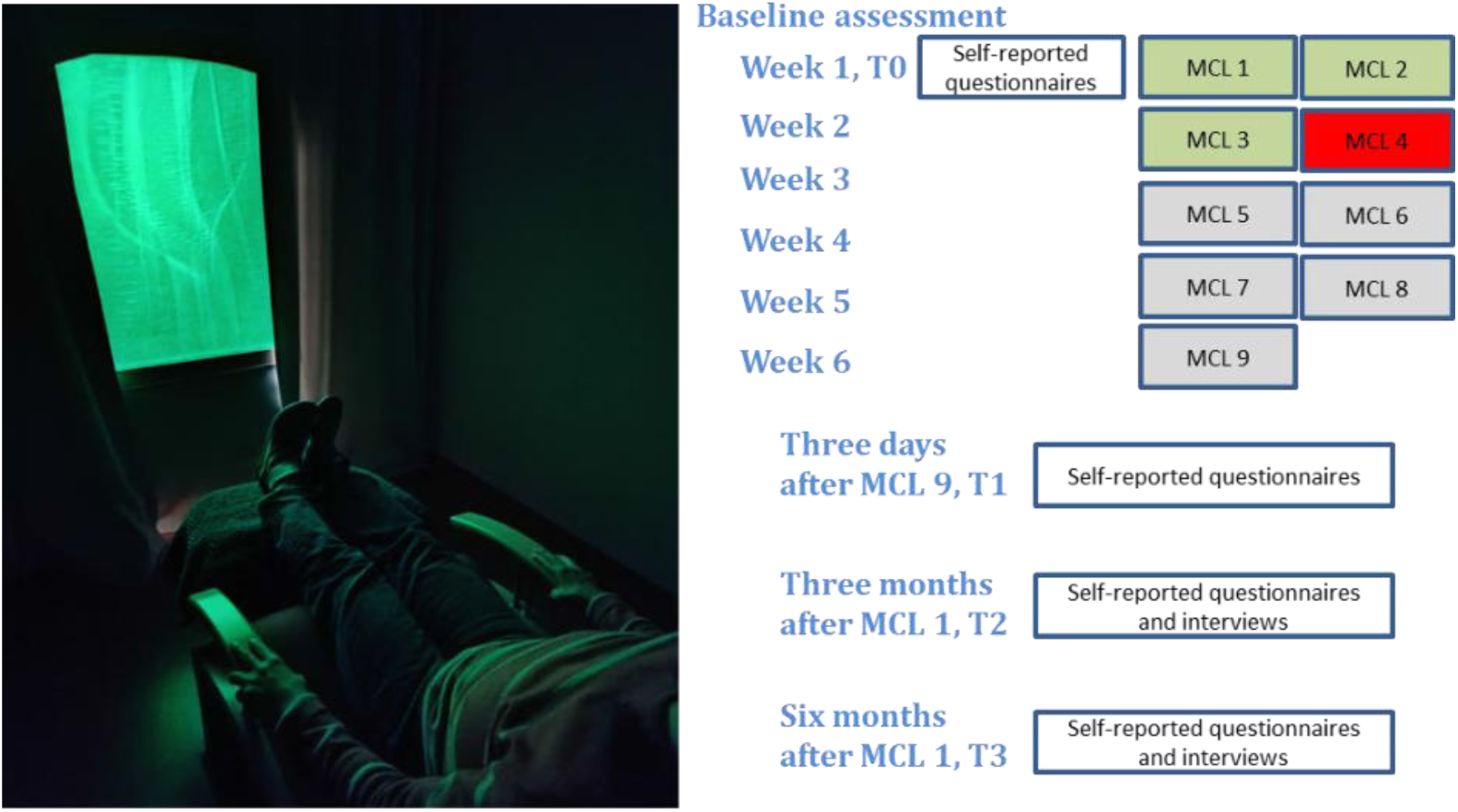
Timeline of the LUCIA study. MCL, metal color light; T0, baseline, day 0, day of first MCL

In each MCL session, the patient and therapist have a preparatory talk of 10-15 minutes before viewing the panel. Viewing typically lasts 5-10 minutes, during which the patient is usually silent. Thereafter, the therapist covers the MCL panel and leaves the room, while the patient is given a 20-minute period of resting on the chair in the darkened room.

A course of MCL therapy consists of nine MCL sessions within five to seven weeks per patient (Figure 1). Patients will be asked to complete self-report questionnaires prior to the first MCL treatment (T0) and three days after the ninth treatment (T1). Three different MCL panels, each approximately one meter in height and 65 centimeters in width, an iron-green panel, a gold-red panel, and a manganese-purple panel, will be used in this study (Figure 2).

**Figure 2.**
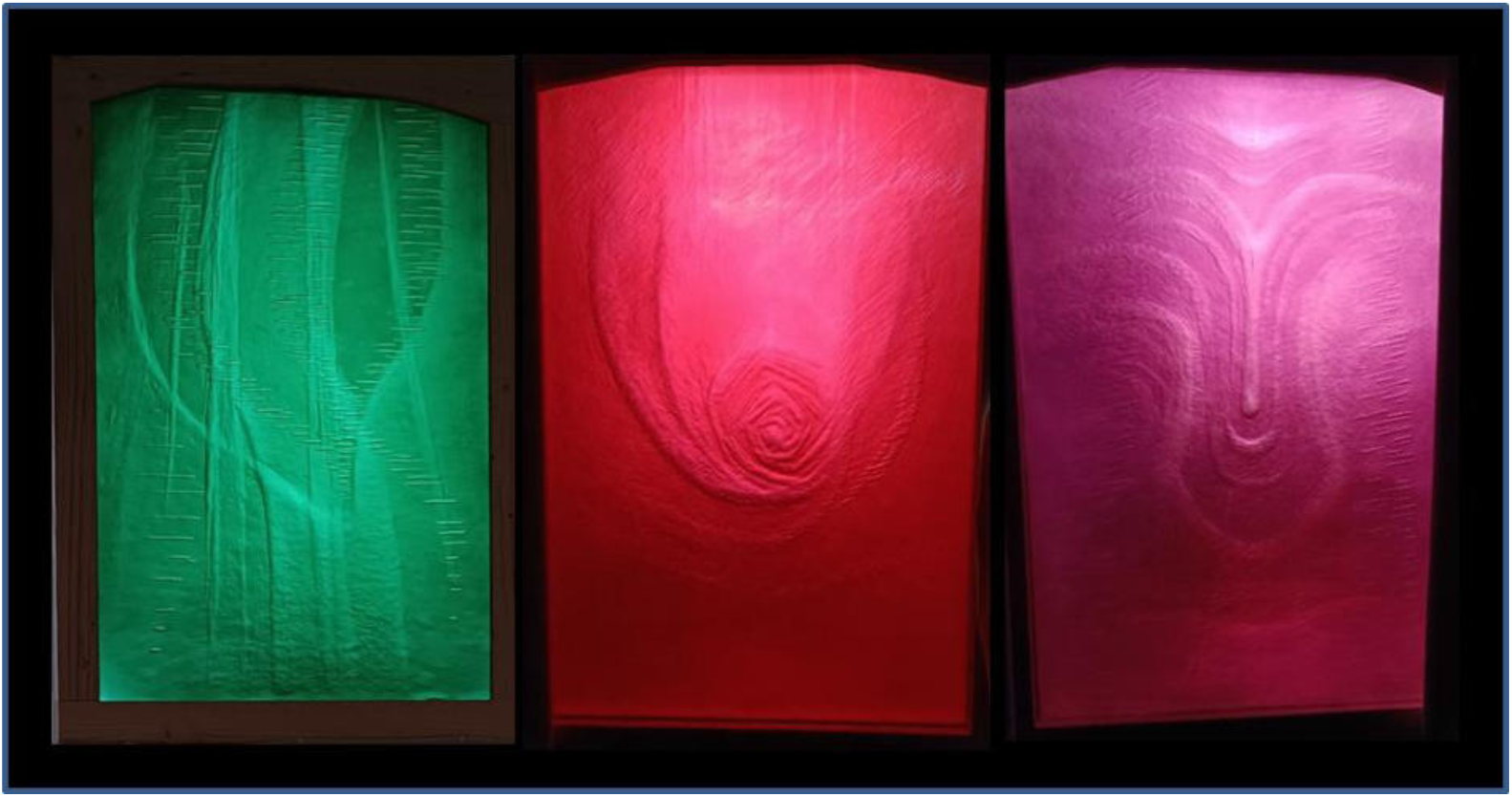
Metal Colored Light panels, iron-green gold-red, manganes-purple.

The intervention is structured in a standardized part and an open section. The first three sessions are done with the iron-green panel, the fourth with the gold-red panel, and the remaining sessions are done individually at the discretion of the therapist (Figure 1). Participants will be interviewed and surveyed at three months (T2) and six months (T3) after the MCL treatment.

### Criteria for discontinuing or modifying allocated interventions

Participants are encouraged to report any discomfort or problems. If a participant does not attend a therapy session, she is contacted by phone, asked how she is feeling, and options are sought how and whether to continue or discontinue the MCL therapy.

### Strategies to improve adherence to interventions and complete follow-up

At the beginning of each MCL therapy session participants are asked about their feeling with the previous session. Their experiences and preferences are discussed as well. The number of sessions each participant actually attended will be recorded and participants will be reminded by phone call to fill in and return self-reported questionnaires. For the follow-up surveys, participants will be contacted by postal mail, by stamped return envelopes enclosed, and by personal telephone calls.

### Relevant concomitant care permitted or prohibited during the trial

Concomitant medications including nutritional supplements, vitamins, and natural remedies will be registered in the case report form. Standard routine care and medication for endocrine treatment should be maintained and will not affect the eligibility of patients to participate in the study. Concomitant sportive activities will be registered and should be maintained as well. This will not affect the eligibility of study participation.

### Outcomes

The primary endpoint is the patient’s self-reported distress using the German version of the National Comprehensive Cancer Network (NCCN) Distress Thermometer [20] before and after nine MCL sessions. Participants will self-rate their experienced distress in the last week on a single 11-point scale ranging from 0 (“No distress”) to 10 (“Extreme distress”). The Distress thermometer is supplemented with a symptoms list including practical, emotional or physical problems such as fatigue, pain, nausea, sleep, or breathing problems. The symptoms that are identified will be evaluated alongside the distress thermometer level but are not part of the primary endpoint.

Secondary endpoints include the self-reported internal coherence measured with the Internal Coherence Scale (ICS) [21] and the self-reported quality of life measured with the European Organization for Research and Treatment of Cancer Questionnaire C30 (EORTC-QLQ-C30) [22]. Equations will be made as described in the ICS and the EORTC-QLQ-C30 manual. The ICS, a short, highly reliable and valid self-report questionnaire, contains two subscales, one with eight items (Inner Coherence and Resilience) and a second subscale (Thermo Coherence) with two items [21]. The EORTC scores range from 0 to 100 and higher scores represent a better self-reported level in the functional dimensions but a higher degree of symptom burden.

Baseline information on diagnosis, such as histology, tumor classification and previous treatments will be collected from the clinical interview at study inclusion and the participants’ medical records. All participants will be asked to answer all questionnaires at baseline (T0) and after the treatment at T1, (Figure 1). The same self-reported questionnaires will be used to monitor changes at the follow-up assessments at T2 and T3. The courses of stress perception and quality of life of participants of the LUCIA-study will be compared to those of other breast cancer patients of the network oncology [23], matched for age, breast cancer type, and oncological treatment.

### Implementation

All patients who give their written consent to participate and fulfil the inclusion criteria will be enrolled in this study and will be documented by the investigator. All patient data is handled in accordance with the General Data Protection Regulation (GDPR). All data from the participants will be maintained confidentially before, during, and after the trial and is stored securely at the study site, with limited access. The study design and participant timeline is depicted in Figure 1.

### Sample size

The expected longitudinal differences between the means of the outcomes assessed will be estimated based on our previously published breast cancer studies [24,25]. For sample size calculation, assuming a power of 90%, a significance level of 5% and a small to medium effect size (d > 0.2), a total of 50 patients would be required to confirm a statistically significant treatment effect according to Schoenfeld et al. [26]. Assuming a dropout rate of 20%, 60 patients need to be recruited for this study.

### Recruitment

Female breast cancer patients who have been treated at the GKH Breast Cancer Center are contacted and invited to participate in this study. A clinical interview is scheduled with interested and eligible patients. After obtaining written informed consent questionnaires will be given to participants.

### Statistical methods

All statistical analyses will be performed using Excel 2010 (Microsoft, USA) and the software R (R Version 4.0.5) [27]. Baseline data will be summarized using descriptive statistics. Continuous variables will be described as means with standard deviation, median with interquartile range (IQR); categorical variables summarized as frequencies and percentages. Student’s t-tests will be applied, to detect group and longitudinal differences, p-values < 0.05 are considered to be significant. For characterization of group differences, Pearson’s Chi-squared tests with Yates’ continuity correction will be performed. To identify influencing factors and to address potential sources of bias and potential confounders, adjusted multivariable linear regression analyses will be performed. In order to yield reliable model results, stepwise regression selections will be performed and models with high adjusted R^2^ will be chosen. According to Cohen’s interpretation [28] R^2^ values between 13% to 25% indicate medium and R^2^ values 26% or above indicate high effect sizes. In addition to statistical significance, we will report descriptive statistics and effect size estimates. Multilevel modeling can be used for subgroup analyses and to account for confounding variables. Confounding variables (age, body mass index, tumor stage, hormonal status, endocrine treatment, concomitant medication, and the respective outcome values at baseline) will be considered. Furthermore, adjusted multivariable linear regression analyses will be performed, to address potential confounders and influencing factors. Subgroup analyses will be performed regarding possible confounders such as concurrent endocrine treatment and concomitant medications for symptom relief.

### Methods in analysis to handle protocol non-adherence and any statistical methods to handle missing data

Patients who are unable to participate in the MCL treatment or withdraw consent to participate in the study will be excluded and not replaced. If individual sessions cannot be attended, this will be noted and the surveys will continue as planned. Regarding the evaluation of questionnaires we follow the EORTC-QLQ-C30 scoring manual on calculation and handling of missing data. We do not plan to use imputation when reporting outcome data. Per-protocol as well as intention-to treat analyses will be performed. An interim analysis will be performed after 20 participants have received the MCL treatment.

### Adverse event reporting and harms

During each MCL therapy session information on the occurrence of adverse events is regularly retrieved and documented. Adverse events related to the MCL treatment during the six weeks will be recorded by the coordinating investigator.

### Patient and Public Involvement

Patients are involved in the selection of MCL panels during MCL therapy sessions (patient decision making). Participants in the study will be involved in the focus of the development of research questions, and their suggestions and objections will be taken into account.

### Dissemination plans

The study results will be presented in conferences and symposia and submitted for publication in relevant peer-reviewed medical journals. Parts from this study will also be included in a doctoral thesis.

## Discussion

In the proposed study project, the effects of metal color light therapy on the perception of stress and breast cancer patients’ well-being will be evaluated. The expected therapeutic effects can unfold on many levels. These effects may range from relaxation and mood enhancement through warming, deepening of breathing, and specific organ effects, to support in coping with illness and or biographical crises. Bright light therapy is effective in the treatment of depression and delayed or advanced sleep phase syndromes [29]. Scientific, systematic investigations and studies on the application of light therapy in people with or after cancer have scarcely been carried out so far [13,12]. The effects of light therapy on fatigue and sleep disturbances in breast cancer survivors have been evaluated in a controlled trial and preliminary results have been published [30]. MCL can be used for people with chronic diseases of the intestines, respiratory and cardiovascular systems, cancer, and also for states of exhaustion [17]. Indications for cancer are varied and include relief from symptoms of respiratory problems, pain, exhaustion, depression and other stress and outpatient MCL can provide a supportive service for coping with and overcoming the disease [17]. Evaluating the distress experienced by breast cancer patients after their primary standard care as it has been the focus of the present study may help to determine which stress symptoms can be influenced by MCL. The purpose of this study is to determine whether MCL is an appropriate therapeutic approach for improving well-being and reducing stress in breast cancer patients. In order to generate hypotheses for follow-up studies, qualitative evaluations regarding the described impressions and feelings of the MCL participants can be considered in the future.

### Trial status

The current protocol version is 1.0, dated from 10. August 2023. The trial is ongoing and currently enrolling. The first participant was enrolled on 06 September 2023 and recruitment is expected to be completed in the 2nd quarter of 2025 with the follow-up completed by the end of 2025.

## Data Availability

Any material required to support the protocol can be supplied on reasonable request.

## Declarations

### Patient and public involvement

Patients and/or the public were not involved in the design, or conduct, or reporting, or dissemination plans of this research.

## Acknowledgements

We would like to thank Friedlinde Meyer for her supportive discussions regarding the establishment of the MCL therapy and the design of the study. Friedlinde Meier was deeply committed to Metal Color Light therapy. She helped to initiate and accompanied this study with all her strength. Unfortunately she passed away on May 6, 2024. We thank her from the bottom of our hearts. We would also like to thank Irena Cop and all the other members of the medical staff at the Havelhöhe Hospital for their support in this work.

## Ethics approval and consent to participate

This study complies with the principles laid down in the Declaration of Helsinki and has been approved by the Ethics Committee of the Berlin Medical Association (Ethik-Kommission der Ärztekammer Berlin) under the number Eth-27/10 and Amendment 7 on February 2, 2023. This trial was registered at the German Clinical Trials Register (Trial registration number DRKS00013335 on 27/11/2017. Written informed consent will be obtained from all participants prior to study enrollment. All substantial protocol deviations or modifications will be communicated to the Ethics Committee and German Clinical Trials Register.

## Availability of data and materials

Any material required to support the protocol can be supplied on reasonable request.

## Consent for publication

Model consent forms will be provided on request.

## Author Statement

SLO conceived the study, initiated the study design, developed the methodology, was responsible for data curation, formal analysis, validation, visualization, wrote the first draft of the manuscript and reviewed it. JG conceived the study, initiated the study design, developed the methodology, was responsible for validation, writing, reviewing, and recruitment of participants. MG conceived the study, initiated the study design, developed the methodology, was responsible for validation, visualization, writing, reviewing and implemented the MCL protocol. GG conceived the study, initiated the study design, developed the methodology, and was responsible for validation, visualization, writing, reviewing, and recruitment of participants. LF conceived the study, initiated the study design, developed the methodology, and was responsible for validation, visualization, writing, reviewing, and recruitment of participants. SJ conceived the study, initiated the study design, developed the methodology, and was responsible for validation, writing and reviewing. FS is the principal investigator of the study, he conceived the study, initiated the study design, developed the methodology, and was responsible for funding acquisition, validation, visualization, writing, reviewing and project administration. AT is the scientific coordinator of the study, she conceived the study, initiated the study design, developed the methodology, was responsible for data curation, formal analysis, validation, visualization, writing, and reviewing. All authors commented on drafts of the manuscript and read and approved the final manuscript.

## References

1. Bray F, Ferlay J, Soerjomataram I, Siegel RL, Torre LA, Jemal A (2018) Global cancer statistics 2018: GLOBOCAN estimates of incidence and mortality worldwide for 36 cancers in 185 countries. CA: a cancer journal for clinicians 68 (6):394–424. doi:10.3322/caac.21492

2. Fabi A, Falcicchio C, Giannarelli D, Maggi G, Cognetti F, Pugliese P (2017) The course of cancer related fatigue up to ten years in early breast cancer patients: What impact in clinical practice? Breast (Edinburgh, Scotland) 34:44–52. doi:10.1016/j.breast.2017.04.012

3. Abrahams HJG, Gielissen MFM, Verhagen C, Knoop H (2018) The relationship of fatigue in breast cancer survivors with quality of life and factors to address in psychological interventions: A systematic review. Clinical psychology review 63:1–11. doi:10.1016/j.cpr.2018.05.004

4. Bower JE (2014) Cancer-related fatigue--mechanisms, risk factors, and treatments. Nature reviews Clinical oncology 11 (10):597–609. doi:10.1038/nrclinonc.2014.127

5. Greenlee H, DuPont-Reyes MJ, Balneaves LG, Carlson LE, Cohen MR, Deng G, Johnson JA, Mumber M, Seely D, Zick SM, Boyce LM, Tripathy D (2017) Clinical practice guidelines on the evidence-based use of integrative therapies during and after breast cancer treatment. CA: a cancer journal for clinicians 67 (3):194–232. doi:10.3322/caac.21397

6. Lipsett A, Barrett S, Haruna F, Mustian K, O’Donovan A (2017) The impact of exercise during adjuvant radiotherapy for breast cancer on fatigue and quality of life: A systematic review and meta-analysis. Breast (Edinburgh, Scotland) 32:144–155. doi:10.1016/j.breast.2017.02.002

7. Bradt J, Dileo C, Magill L, Teague A (2016) Music interventions for improving psychological and physical outcomes in cancer patients. The Cochrane database of systematic reviews (8):CD006911. doi:10.1002/14651858.CD006911.pub3

8. Schad F, Rieser T, Becker S, Groß J, Matthes H, Oei SL, Thronicke A (2023) Efficacy of Tango Argentino for Cancer-Associated Fatigue and Quality of Life in Breast Cancer Survivors: A Randomized Controlled Trial. Cancers (Basel) 15 (11):2920

9. Galassi F, Merizzi A, D’Amen B, Santini S (2022) Creativity and art therapies to promote healthy aging: A scoping review. Frontiers in psychology 13:906191. doi:10.3389/fpsyg.2022.906191

10. Abbing A, Baars EW, de Sonneville L, Ponstein AS, Swaab H (2019) The Effectiveness of Art Therapy for Anxiety in Adult Women: A Randomized Controlled Trial. Frontiers in psychology 10:1203. doi:10.3389/fpsyg.2019.01203

11. Tang Y, Fu F, Gao H, Shen L, Chi I, Bai Z (2019) Art therapy for anxiety, depression, and fatigue in females with breast cancer: A systematic review. Journal of psychosocial oncology 37 (1):79–95. doi:10.1080/07347332.2018.1506855

12. Robijns J, Censabella S, Bulens P, Maes A, Mebis J (2017) The use of low-level light therapy in supportive care for patients with breast cancer: review of the literature. Lasers Med Sci 32 (1):229–242. doi:10.1007/s10103-016-2056-y

13. Mahmood D, Ahmad A, Sharif F, Arslan SA (2022) Clinical application of low-level laser therapy (Photo-biomodulation therapy) in the management of breast cancer-related lymphedema: a systematic review. BMC cancer 22 (1):937. doi:10.1186/s12885-022-10021-8

14. Sun W, Yan J, Wu J, Ma H (2022) Efficacy and Safety of Light Therapy as a Home Treatment for Motor and Non-Motor Symptoms of Parkinson Disease: A Meta-Analysis. Med Sci Monit 28:e935074. doi:10.12659/msm.935074

15. Kim WS, Calderhead RG (2011) Is light-emitting diode phototherapy (LED-LLLT) really effective? Laser therapy 20 (3):205–215. doi:10.5978/islsm.20.205

16. Emens JS, Burgess HJ (2015) Effect of Light and Melatonin and Other Melatonin Receptor Agonists on Human Circadian Physiology. Sleep Med Clin 10 (4):435–453. doi:10.1016/j.jsmc.2015.08.001

17. Altmaier M (2010) Metallfarblichttherapie, Zur Forschung und Entwicklung einer neuen Therapie auf anthroposophischer Grundlage. . Mayer-Verlag, Germany, Stuttgart

18. Chan AW, Tetzlaff JM, Altman DG, Laupacis A, Gøtzsche PC, Krleža-Jerić K, Hróbjartsson A, Mann H, Dickersin K, Berlin JA, Doré CJ, Parulekar WR, Summerskill WS, Groves T, Schulz KF, Sox HC, Rockhold FW, Rennie D, Moher D (2013) SPIRIT 2013 statement: defining standard protocol items for clinical trials. Annals of internal medicine 158 (3):200–207. doi:10.7326/0003-4819-158-3-201302050-00583

19. Schad F, Axtner J, Happe A, Breitkreuz T, Paxino C, Gutsch J, Matthes B, Debus M, Kroz M, Spahn G, Riess H, von Laue HB, Matthes H (2013) Network Oncology (NO)--a clinical cancer register for health services research and the evaluation of integrative therapeutic interventions in anthroposophic medicine. Forschende Komplementarmedizin 20 (5):353–360. doi:10.1159/000356204

20. Mehnert A, Lehmann C, Cao P, Koch U (2006) [Assessment of psychosocial distress and resources in oncology--a literature review about screening measures and current developments]. Psychother Psychosom Med Psychol 56 (12):462–479. doi:10.1055/s-2006-951828

21. Kroz M, Bussing A, von Laue HB, Reif M, Feder G, Schad F, Girke M, Matthes H (2009) Reliability and validity of a new scale on internal coherence (ICS) of cancer patients. Health and quality of life outcomes 7:59. doi:10.1186/1477-7525-7-59

22. Aaronson NK, Ahmedzai S, Bergman B, Bullinger M, Cull A, Duez NJ, Filiberti A, Flechtner H, Fleishman SB, de Haes JC, et al. (1993) The European Organization for Research and Treatment of Cancer QLQ-C30: a quality-of-life instrument for use in international clinical trials in oncology. Journal of the National Cancer Institute 85 (5):365–376

23. Schad F, Thronicke A, Merkle A, Steele ML, Kroz M, Herbstreit C, Matthes H (2018) Implementation of an Integrative Oncological Concept in the Daily Care of a German Certified Breast Cancer Center. Complement Med Res 25 (2):85–91. doi:10.1159/000478655

24. Oei SL, Thronicke A, Grieb G, Schad F, Groß J (2023) Evaluation of quality of life in breast cancer patients who underwent breast-conserving surgery or mastectomy using real-world data. Breast cancer (Tokyo, Japan) 30 (6):1008–1017. doi:10.1007/s12282-023-01494-x

25. Oei SL, Thronicke A, Matthes H, Schad F (2020) Evaluation of the effects of integrative non-pharmacological interventions on the internal coherence and resilience of breast cancer patients. Supportive care in cancer : official journal of the Multinational Association of Supportive Care in Cancer. doi:10.1007/s00520-020-05617-4

26. Schoenfeld DA (1983) Sample-size formula for the proportional-hazards regression model. Biometrics 39 (2):499–503

27. Team RC (2016) R: A language and environment for statistical computing R Foundation for Statistical Computing , Vienna, Austria

28. Cohen J (1992) A power primer. Psychological bulletin 112 (1):155–159. doi:10.1037//0033-2909.112.1.155

29. Terman M, Terman JS (2005) Light therapy for seasonal and nonseasonal depression: efficacy, protocol, safety, and side effects. CNS spectrums 10 (8):647-663; quiz 672. doi:10.1017/s1092852900019611

30. Wu HS, Gao F, Yan L, Given C (2022) Evaluating chronotypically tailored light therapy for breast cancer survivors: Preliminary findings on fatigue and disrupted sleep. Chronobiol Int 39 (2):221–232. doi:10.1080/07420528.2021.1992419

